# The pneumonia severity index: assessment and comparison to popular machine learning classifiers

**DOI:** 10.1101/2021.12.06.21267390

**Authors:** Dawei Wang, Deanna R. Willis, Yuehwern Yih

**Affiliations:** School of Industrial Engineering, Purdue University, 315 Grant St, West Lafayette, IN 47907, USA; Indiana University School of Medicine, Department of Family Medicine, 1110 W. Michigan St, LO 200, Indianapolis, IN 46202, USA

**Keywords:** Pneumonia Severity Index (PSI), Community Acquired Pneumonia (CAP), machine learning, classification, prediction model

## Abstract

Pneumonia is the top communicable cause of death worldwide. Accurate prognostication of patient severity with Community Acquired Pneumonia (CAP) allows better patient care and hospital management. The Pneumonia Severity Index (PSI) was developed in 1997 as a tool to guide clinical practice by stratifying the severity of patients with CAP. While the PSI has been evaluated against other clinical stratification tools, it has not been evaluated against multiple classic machine learning classifiers in various metrics over large sample size. In this paper, we evaluated and compared the prediction performance of nine classic machine learning classifiers with PSI over 34720 adult (age 18+) patient records collected from 749 hospitals from 2009 to 2018 in the United States on Receiver Operating Characteristic (ROC) Area Under the Curve (AUC) and Average Precision (Precision-Recall AUC). Machine learning classifiers, such as Random Forest, provided a significant improvement (∼29% in PR AUC and ∼5% in ROC AUC) compared to PSI and required only 7 input values (compared to 20 parameters used in PSI). There were also statistically significant differences (p<0.05) between Random Forest and PSI among various races/ethnicities. Because of its ease of use, PSI remains a very strong clinical decision tool, but machine learning classifiers can provide better prediction accuracy performance. Comparing prediction performance across multiple metrics such as PR AUC, instead of ROC AUC alone can provide additional insight.

**Key Messages:** *This work compared the prognostication accuracy performance of patient severity with Community Acquired Pneumonia (CAP) between Pneumonia Severity Index (PSI) and nine machine learning classifiers and found machine learning classifiers provided a significant improvement*.

## 1. INTRODUCTION

Pneumonia is one of the leading causes of death worldwide. Lower respiratory infection is ranked top four leading cause of death globally in 2019 [1]. Furthermore, among communicable diseases, it is the top leading cause of death globally in 2019. Pneumonia is the single largest infectious cause of death in children worldwide. It killed 808,694 children under the age of 5 in 2017, accounting for 15% of all deaths of children under five years old [2]. Between 2015 and 2019, the United States, influenza and pneumonia are ranked in the top eight leading causes of death [3]. Accurate prognostication of patient severity not only allows physicians to determine the most appropriate site of treatment (home vs. hospital) and treatment plan, but also assist the intensity of care management such as admission to an intensive care unit and intravenous antibiotic therapy [4].

During times of shortage in hospital beds, medical supplies, and provider workforce, such as during the COVID-19 pandemic, prognostication of pneumonia patient severity to coordinate proper resources to improve patient outcomes becomes paramount due to the scarcity in resources. During the COVID-19 pandemic in the US, as of April 2021, nearly one in 11 hospitals with intensive care units (ICU) reported that at least 95% of their ICU beds were full and 70% of ICU beds were occupied nationwide [24].

The Pneumonia Severity Index (PSI), developed in 1997, is a common tool to classify the severity of Community-Acquired Pneumonia (CAP) patients using 20 demographic and clinical variables. It was derived from the evaluation of 14,199 adult inpatients with CAP and validated in 1991 in two separate studies using data from 38,039 inpatients and 2,287 inpatients and outpatients, respectively [5]. Since then, because of its prognostic accuracy and ease of use, PSI has become the reference standard for stratifying patient severity with CAP. In 2008, the effectiveness and safety of the PSI was revalidated in multiple studies with a total of 3,949 low-risk patients at 60 sites in 4 countries (US, Canada, France, and Spain) [6–10], and compared the accuracy of PSI with CURB-65 more than five studies with a total of 6,773 patients at 36 sites in 5 countries (US, Australia, Spain, China, and Sweden) [11–15]. Overall, those studies have demonstrated a beneficial impact of the PSI on patient care [4]. Compared to CURB-65, PSI identifies a slightly greater proportion of patients at low risk and has higher discriminatory power for mortality.

Machine learning algorithms, especially deep learning approaches, have evolved a lot over the past decades since PSI was developed in 1997. They have been applied in a variety of healthcare areas, such as: patient risk/disease prediction, treatment suggestion, clinical image recognition, and genomics. Classification problems have been studied widely across various healthcare problems [16]. Specifically, for pneumonia, deep learning has been applied to chest x-ray images to classify pneumonia severity [17, 18]. In 2020, during the COVID-19 pandemic, these approaches were applied to classify COVID-19 cases through x-ray images [19–22]. For example, deep learning approaches have been used to stratify the severity of COVID-19 patients and compared to the performance of the PSI and CURB-65 reference standards [23]. In this study, we are to evaluate and compare the performance of the reference standard PSI with popular machine learning classifiers to understand if the PSI remains a good tool to stratify pneumonia severity nearly 30 years after its development.

## 2. MATERIALS AND METHODS

### 2.1 Data Overview

We used de-identified longitudinal electronic health record (EHR) patient data that was captured in the Cerner database. This dataset contains 1,023,814 adult (age greater than 18 years of age) patient records with pneumonia as principal diagnosis collected from 749 hospitals in the United States. Among those patients from 2009 to 2018, 34,720 records contained all laboratory test results and clinical values required for calculation of the PSI (listed in Table 1). A widely cited reference article describing the input parameters of the PSI was used to define required variables [5]. Encounters with the ICD-9 and ICD-10 diagnosis codes of CAP were used to identify cases of CAP. These included the following ICD-9 and ICD-10 codes: 480.*, 486.*, 487.*, J13, J14, J15.211, J15.3, J15.4, J15.7, J15.9, J16.0, J16.8, J18.0, J18.1, J18.8, J18.9. Death cases were counted upon patient discharge.

**Table 1:** Cerner data overview of Community-Acquired Pneumonia Severity Index (PSI) for Adults: total number and in-hospital mortality of each group by demographic, comorbidity, physical examination findings, laboratory, and radiographic findings

|  | Death | Total | In-Hospital Mortality (%) |
| --- | --- | --- | --- |
| **Overall** | 1710 | 34720 | 4.92 |
| **Sex** |  |  |  |
| Male (0 points) | 900 | 16548 | 5.43 |
| Female (-10 points) | 810 | 18172 | 4.45 |
| **Demographic** |  |  |  |
| Nursing home resident (10 points) | 182 | 1355 | 13.43 |
| **Comorbid illnesses** |  |  |  |
| Neoplastic disease (30 points) | 358 | 3320 | 10.78 |
| Liver disease (20 points) | 75 | 762 | 9.84 |
| Congestive heart failure (10 points) | 182 | 2283 | 7.97 |
| Cerebrovascular disease (10 points) | 14 | 230 | 6.08 |
| Renal disease (10 points) | 416 | 6088 | 6.83 |
| **Physical examination findings** |  |  |  |
| Respiratory rate $\leq 30$/min (20 points) | 277 | 2561 | 10.81 |
| Systolic blood pressure $< 90$ mmHg (20 points) | 150 | 1319 | 11.37 |
| Temperature $< 35^\circ\text{C}$ or $\geq 40^\circ\text{C}$ (15 points) | 36 | 236 | 15.25 |
| Pulse $\leq 125$/min (10 points) | 138 | 1644 | 8.39 |
| **Laboratory and radiographic findings** |  |  |  |
| Arterial pH $< 7.35$ (30 points) | 491 | 2783 | 17.64 |
| Blood urea nitrogen $\geq 30$ mg/dL (20 points) | 788 | 8160 | 9.65 |
| Sodium $< 130$ mmol/L (20 points) | 175 | 2035 | 8.59 |
| Glucose $\geq 250$ mg/dL (10 points) | 127 | 2140 | 5.93 |
| Hematocrit $< 30\%$ (10 points) | 394 | 4823 | 8.16 |
| Oxygen saturation $< 90\%$ (10 points) | 225 | 2748 | 8.18 |
| Pleural effusion (10 points) | 230 | 2554 | 9.00 |

### 2.2 Preprocessing

The arterial blood pH test, measuring the acidity (pH) of the blood, is usually ordered based on clinical assessment when a patient is viewed to be critically ill and may require the assistance of mechanical ventilation. Not all pneumonia patients had this test. This means that this information was not missing at random. Patients who had this information might be more severe than those who didn’t. This makes the most likely clinical reason why the value is missing being that the treating clinician may not have ordered an arterial blood gas because the patient was not so ill as to require one. Therefore, patients without this information were assumed to be within the normal range. In cases where the patients didn’t have this test performed, two approaches were used to fill in the missing value for the arterial pH test. One method named “mean value imputation” (MVI) [26] filled the missing arterial pH value with the mean value of all other patients’ pH test results in this dataset. The mean value (value = 7.38) is within the normal range (value ≥ 7.35). The other method was to convert all Arterial pH values to 0 and 1 where “1” indicates “out of normal range” (value <7.35), and “0” indicates “normal” for all other arterial pH values, including no test. The arterial pH severity threshold 7.35 was inferred from the PSI guideline (shown in Table 1). Since the prediction performance of classifiers may benefit differently from each preprocessing approach, results of both approaches are shown in the result section.

### 2.3 Performance Measurements

To compare the performance of PSI and machine learning based classifiers, we reviewed the commonly used measurements in both medical and machine learning fields. The following confusion matrix explains the cases of True Positive (TP), False Positive (FP), True Negative (TN), and False Negative (FN). The sum of TP and FN is the total number of positive cases (P) while the sum of TN and FP is the total number of the negative cases (N).

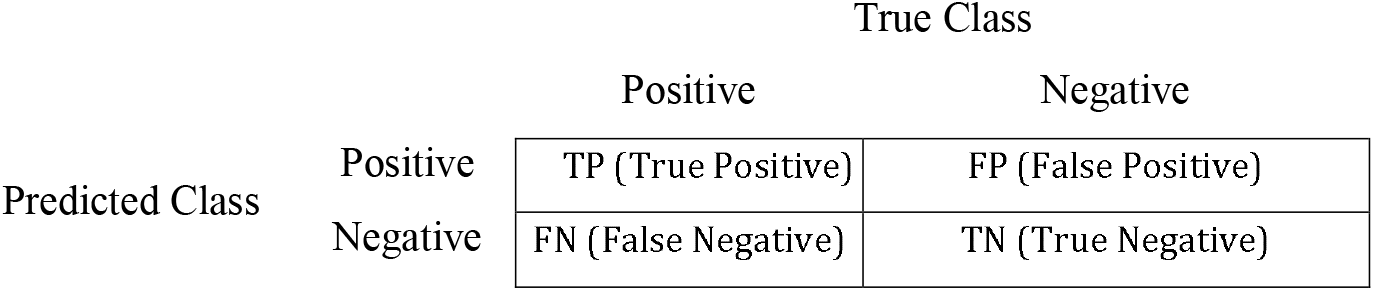

|  |  | True Class |  |
| --- | --- | --- | --- |
|  |  | Positive | Negative |
| Predicted Class | Positive | TP (True Positive) | FP (False Positive) |
|  | Negative | FN (False Negative) | TN (True Negative) |

Precision, sensitivity (recall, True Positive Rate (TPR)), specificity (True Negative Rate (TNR)), False Positive Rate (FPR), and Average Precision (AP) are defined as:

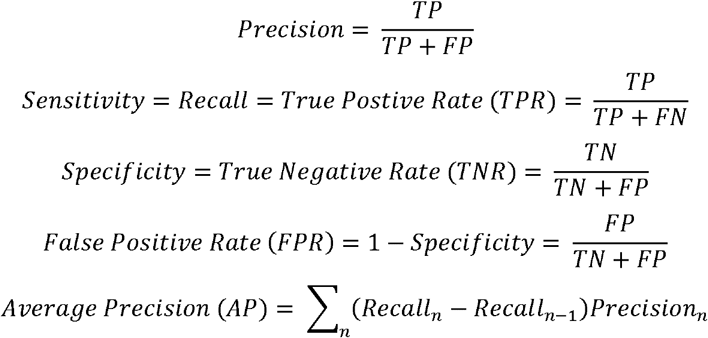

Where Precision_n_ and Recall_n_ are the precision and recall at nth threshold.

In the medical field, specificity, sensitivity, and the ROC-AUC score (the area under the Receiver Operating Characteristic curve) are commonly used. Specificity tells us how well the test performs to identify negative results out of all people that do not have the disease. Sensitivity tells us how well the predictive model performs to identify positive results out of all people that are truly positive. The ROC-AUC score, determined by the area under the ROC curve plotting True Positive Rate (TPR) vs. False Positive Rate (FPR) to measure how well the model distinguishes classes (positive cases vs. negative cases in this scenario).

In machine learning, it is common to look at precision and recall to evaluate the prediction accuracy performance of the models. Precision tells us how many people predicted as positive are true positives. Recall is calculated the same as sensitivity. A precision-recall (PR) curve focuses on the performance of a classifier on the positive class (death or patient with disease) only. In our case, with death cases as positive class, the data is imbalanced with mortality rate=4.92%. With a dataset consisting of very few positive cases (death cases), using the ROC curve to estimate the classification performance could lead to deceptive interpretation of specificity. On the other hand, the PR curve can provide more reliable estimation of prediction accuracy since it evaluates the fraction of true positives among positive predictions [27]. It is more sensitive to the improvements for the positive class. Average Precision (AP) or PR AUC summarizes a PR curve as the weighted mean of precisions achieved at each threshold, with the increase in recall from the previous threshold used as the weight [28].

Although ROC AUC is widely used in evaluating the performance of models in medical field, it weights false positives and false negatives equally. This may not reflect the proper tradeoffs between these two measures in the clinical setting, where false negatives may be much worse than false positives. However, we recognize that a model producing 0% false negatives could mean it classifies all cases to be positive so there are no “misses”, and it doesn’t have any distinguishing power. This is the main reason ROC AUC is a popular choice although it’s not perfect. ROC AUC is used in this paper as a comparison to the performance of the reference standard (PSI) as it was reported in their original paper [5]. As discussed earlier, AP is more suitable to reflect the improvements for positive class.

### 2.4 Experiments

Nine popular machine learning classifiers were evaluated and compared with the PSI. The tested classifiers were: Decision Tree, Random Forest, Gaussian Naive Bayes, Logistic Regression, Linear Discriminant Analysis, Stochastic Gradient Descent, Support Vector Machine (SVM), K-Nearest Neighbors, and Multi-Layer Perceptron. Scikit-learn [25] machine learning package in Python was used for implementation and analyses. Some classifiers are known to perform better with scaled or weighted datasets (e.g.: SVM). Three sets of experiments for each classifier were conducted with default, scaled, and weighted class setting. Robust Scaler from scikit-learn was used for scaling. Area under the receiver operating characteristic curve (ROC-AUC) score and Average Precision (AP) were used as the measurement of performance. Each experiment for the classic classifiers was conducted using random permutation cross validation repeated 5 times with the same segmentation across all tests so that none of the tests would benefit from a particular split. The training data contained 80% of the data with the rest 20% used for testing each time. Each measurement was averaged over 5 tests.

The performance of the 9 classifiers with Top-K features were also compared. Univariate feature selection was used with ANOVA f-test.

## 3. RESULTS AND DISCUSSION

### 3.1 ROC-AUC score

In this section, ROC-AUC score among PSI and all classifiers are compared since the PSI performance is measured in ROC-AUC score in the original study.

The performance of PSI was measured with optimal threshold to maximize the ROC-AUC score. The performance of all machine learning classifiers was shown with their default configuration from the scikit-learn library [25], classes weight being balanced, and input data being scaled. Using different preprocessing approaches of filling the missing pH values, the performance of the classifiers varied. With arterial pH missing values filled with the mean (7.38), two classifiers (scores in bold), Random Forest and Multi-Layer Perceptron (MLP) performed 5.19% and 1.28% better than baseline PSI (listed in Table 2). With arterial pH value filled with “1” if arterial pH< 7.35 and “0” otherwise or missing, five classifiers (as shown in Table 2 scores in bold) performed better than baseline PSI. Logistic Regression and Multi-Layer Perceptron (MLP), performed the best with 5.26% improvement over the PSI. The best performance for MLP was achieved with 1 layer and 5 nodes. More layers or nodes did not improve the performance in terms of ROC-AUC score.

**Table 2:** Prediction accuracy (ROC AUC score) of PSI and classifiers

| Method | Arterial pH missing values = mean (7.38) |  |  |  | Arterial pH = “1” if pH < 7.35, all others “0” |  |  |  |
| --- | --- | --- | --- | --- | --- | --- | --- | --- |
|  | Default | Weighted | Scaled | Improvement (%) | Default | Weighted | Scaled | Improvement (%) |
| PSI | 0.77 | - | - | - | 0.76 | - | - | - |
| Random Forest | **0.81** | **0.81** | **0.80** | **5.19** | **0.78** | **0.78** | **0.78** | **2.63** |
| Multi-Layer Perceptron | 0.75 | - | **0.78** | **1.28** | **0.79** | - | **0.80** | **5.26** |
| Logistic Regression | 0.76 | - | 0.76 | - | **0.79** | - | **0.80** | **5.26** |
| Linear Discriminant Analysis | 0.75 | - | 0.75 | - | **0.79** | - | **0.79** | **3.94** |
| Gaussian Naive Bayes | 0.77 | - | 0.77 | - | **0.77** | - | **0.77** | **1.32** |
| Stochastic Gradient Descent | 0.69 | 0.73 | 0.57 | - | 0.74 | 0.75 | 0.58 | - |
| Support Vector Machine | 0.60 | 0.73 | 0.61 | - | 0.55 | 0.73 | 0.65 | - |
| K-Nearest Neighbors | 0.57 | - | 0.59 | - | 0.56 | - | 0.69 | - |
| Decision Tree | 0.57 | 0.55 | 0.56 | - | 0.56 | 0.54 | 0.56 | - |

### 3.2 Feature Selection

Univariate feature selection was conducted using analysis of variance (ANOVA) f-test to understand if we could achieve similar performance with less clinical data compared to PSI. Feature importance with two approaches to handle arterial pH missing values are shown in Fig. 1 and their ROC-AUC scores of classifiers with different K features are shown in Fig. 2. For comparison purpose, PSI performance is shown as a reference line in Fig 2, noting that PSI always requires all features and doesn’t have an option for feature reduction. With arterial pH missing values filled with mean shown in Fig 2(a), Gauss Naïve Bayes (Gauss) and Random Forest (Forest) performed better than the PSI with as few as the top 6 or 7 features accordingly. With arterial pH filled with “1” or “0” based on the severity threshold value of 7.35 in Fig 2(b), Multi-Layer Perceptron (MLP), Gauss Naïve Bayes, Linear Discriminant Analysis, and Logistic Regression (Log) performed better than PSI, with as few as 2 nodes and the top 6 to 7 features accordingly. Blood urea nitrogen and arterial pH are the top two features to predict in-hospital mortality (shown in Fig. 1).

**Figure 1:**
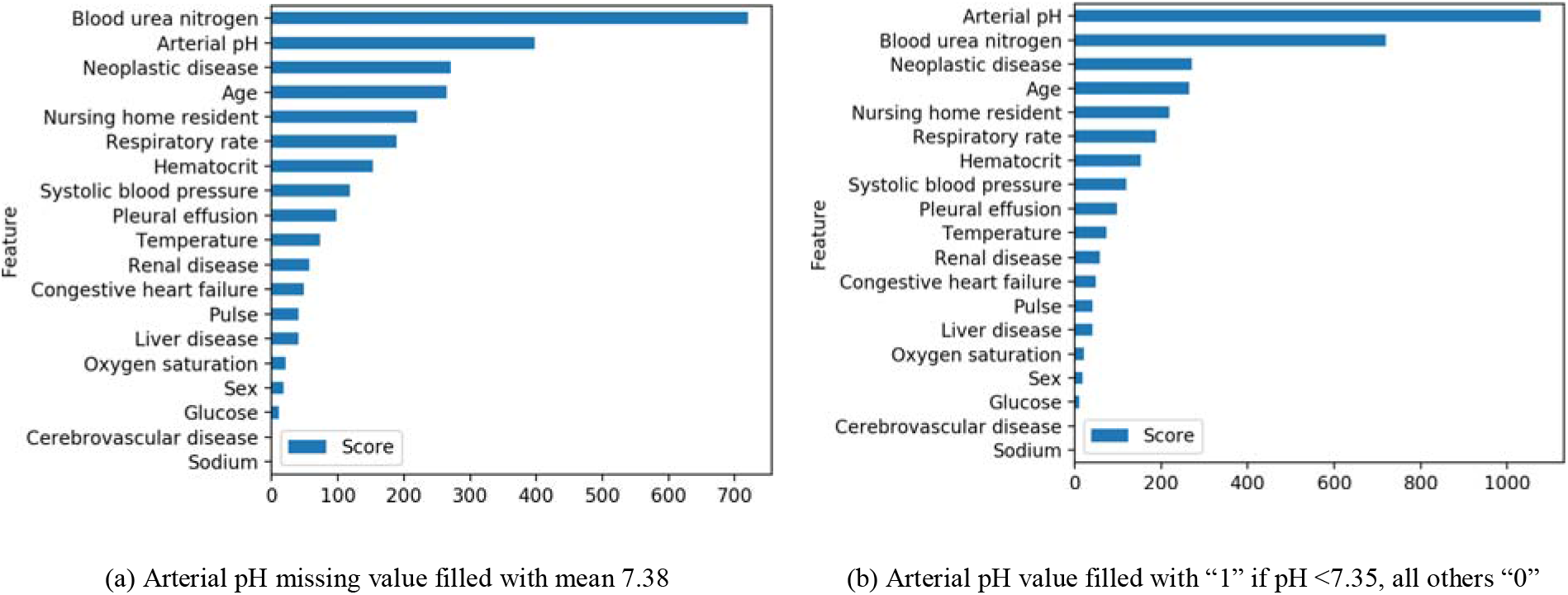
Feature importance of predicting mortality using ANOVA f-test

**Figure 2:**
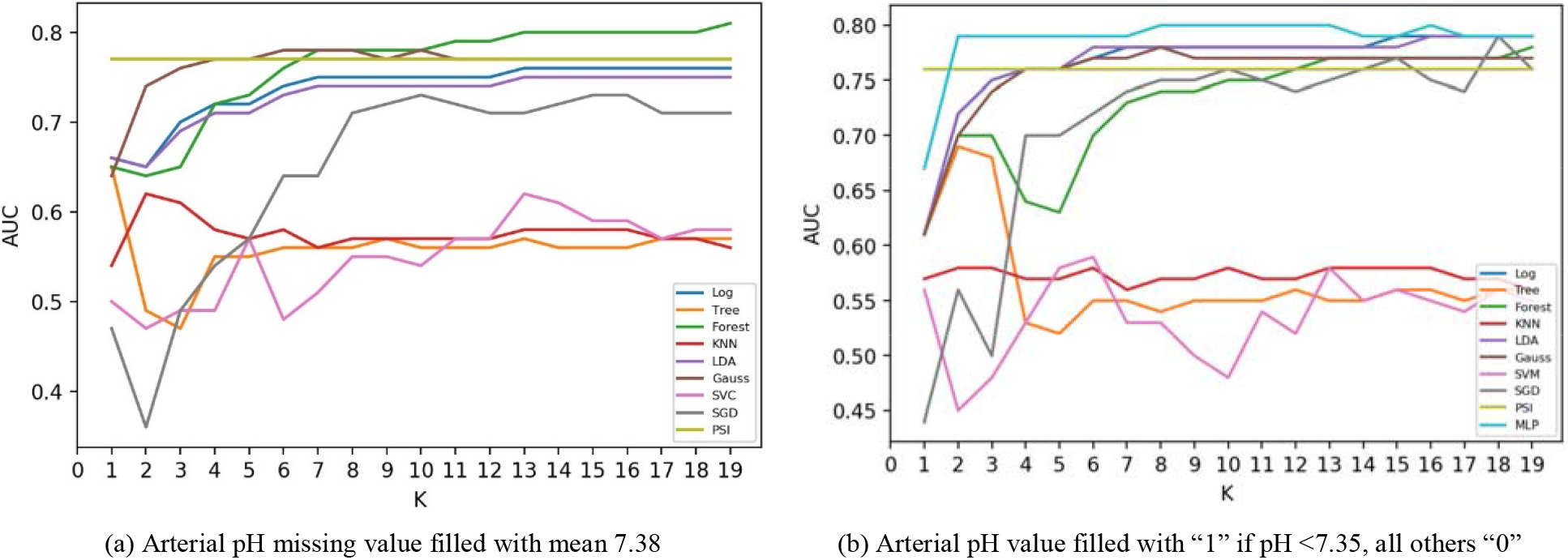
In-hospital mortality prediction performance of traditional classification algorithm with Top K features vs. PSI in ROC-AUC score

### 3.3 ROC-AUC Score Among Patient Races

Since Random Forest (default setting from scikit-learn [25], with missing arterial pH values filled with mean=7.38) performed the best in terms of ROC-AUC score among all machine learning classifiers, it was chosen to compare to the PSI to investigate if any performance variance exists between Caucasians and African Americans. The dataset contains 28222 Caucasians and 2951 African Americans. The datasets for other racial groups are too small for comparison: 726 Native Americans, 539 Hispanics, 501 Asians, 80 Biracials, 13 Pacific islanders, 9 Mid-Eastern Indians and 3 Asian/Pacific Islanders.

Instead of random permutation cross-validation repeated 5 times by sampling 80% of the data for training and 20% for testing as the rest of the paper, 90% of the data was sampled for training and 10% for testing for Random Forest (balanced class weight, number of estimators=1000). Since both 80% training sample size for African Americans and 5 data points for paired t-test were too small. The same ten testing datasets were used for PSI for comparison. A paired t-test of ROC AUC scores from the 10 tests was conducted between PSI and Random Forest among each racial/ethnic group. The prediction accuracy of both the PSI and random forest, in terms of ROC-AUC score, varied based on patient race/ethnicity. Overall, random forest performed better than PSI among all patient races/ethnicities. The ROC AUC score for the Random Forest was statistically significantly higher than that of PSI. The mean of the paired differences of Random Forest and PSI for Caucasians was 0.04 (p <0.001). The mean of the paired differences of Random Forest and PSI for African Americans was 0.03 (p<0.005). Patient race/ethnicity was not highly correlated (close to 0) compared to other clinical and demographic variables, based on analyses using the ANOVA f-test.

Racial and ethnic disparities in risk for pneumonia severity and mortality have been shown to be related to a variety of factors related to processes and systems of care. For example, compared to whites, African Americans and Hispanics have previously been shown to be less likely to receive pneumococcal vaccination, smoking cessation counselling, and influenza vaccinations. In addition, racial/ethnic disparities in pneumonia care occur both within and between hospitals, but that between-hospital disparities are more consistent across measures and are often of greater magnitude [29]. These systemic and system factors necessitate further exploration to understand how PSI and classifiers underperforms in some racial/ethnic groups.

### 3.4 PR AUC Score (Average Precision)

In this section, average precision among PSI and all classifiers are compared.

To compare mortality prediction in terms of PR AUC score, the results for average precision among PSI and classifiers of the similar experiments and settings to the previous section are shown in Tables 3. The performance in these settings have not been optimized by average precision estimators, with the estimator and parameters unchanged from previous sets of experiments performed in the section for ROC-AUC score. As mentioned in the performance measurement section, PR Curve is more sensitive to improvements over positive cases. We could see that the improvements of the performances for machine learning classifiers compared to PSI were larger in terms of average precision (∼29%) compared to those in terms of ROC AUC (∼5%) shown in Table 2. Using different preprocessing approaches of filling the missing pH values, the performance of the classifiers varied. With arterial pH missing values filled with the mean (7.38), four classifiers (scores in bold) performed better than baseline PSI (listed in Table 3). Among those four classifiers, Random Forest (0.22) achieved the best with 29.41% improvement compared PSI (0.17). With arterial pH value filled with “1” if arterial pH< 7.35 and “0” otherwise or missing, four classifiers performed better than baseline PSI. Among those four classifiers, Multi-Layer Perceptron (MLP) performed the best with 23.52% improvement over the PSI. The best performance for MLP was achieved with 1 layer and 5 nodes. More layers or nodes did not improve the performance in terms of average precision.

**Table 3:** Average Precision of PSI and classifiers

| Method | Arterial pH missing values = mean (7.38) |  |  |  | Arterial pH = “1” if pH < 7.35, all others “0” |  |  |  |
| --- | --- | --- | --- | --- | --- | --- | --- | --- |
|  | Default | Weighted | Scaled | Improvement (%) | Default | Weighted | Scaled | Improvement (%) |
| PSI | 0.17 | - | - | - | 0.17 | - | - | - |
| Random Forest | **0.22** | **0.19** | **0.21** | **29.41** | **0.20** | 0.17 | **0.20** | **17.65** |
| Multi-Layer Perceptron | 0.16 | - | **0.20** | **17.65** | **0.20** | - | **0.21** | **23.52** |
| Linear Discriminant Analysis | **0.19** | - | **0.19** | **11.76** | **0.19** | - | **0.19** | **11.76** |
| Logistic Regression | **0.18** | - | **0.18** | **5.88** | **0.20** | - | **0.20** | **17.65** |
| Gaussian Naive Bayes | 0.15 | - | 0.15 | - | 0.14 | - | 0.14 | - |
| Stochastic Gradient Descent | 0.13 | 0.13 | 0.12 | - | 0.16 |  | 0.10 | - |
| Support Vector Machine | 0.09 | 0.14 | 0.12 | - | 0.08 |  | 0.14 | - |
| K-Nearest Neighbors | 0.07 | - | 0.07 | - | 0.07 | - | 0.08 | - |
| Decision Tree | 0.07 | 0.06 | 0.07 | - | 0.07 | 0.06 | 0.07 | - |

Overall, the PSI effectively turns continuous clinical laboratory and biometric values into categorical variables for weighting in the PSI tool. Machine learning classifiers work better than PSI because the machine learning classifiers takes raw laboratory testing and exam results as continuous-variable model inputs. Thus, machine learning classifiers can potentially be more precise on determining the severity score to stratify the severity of the patients. While on the other hand, PSI has prefixed threshold for each test result and score for each result that passes the threshold.

## 4. CONCLUSIONS

We compared the prediction performance of the PSI with 9 classic machine learning classifiers with 20 clinical and demographic variables. We found some classic machine learning classifiers could potentially perform better than the PSI in terms of ROC-AUC and PR-AUC scores. Among all 9 machine learning classifiers, Random Forest can improve the prediction accuracy by as much as 29.41% in terms of the PR-AUC score and 5.19% in terms of the ROC-AUC score compared to PSI. As electronic health records develop automated real-time decision support tools to predict pneumonia severity, Random Forest may reduce missed cases of severe pneumonia and potentially save lives. It could perform better than PSI with as few as just 7 input values. This implies that with machine learning based classifiers, we could achieve similar prediction performance levels with fewer clinical tests and data, which could lead to reduction in wait time (delay) in triaging patients to provide adequate level of care promptly. It’s noted that the performance of PSI and Random Forest varied among patients of different ethnic groups, with Random Forest performing statistically significant (p<0.05) better among Caucasians and African Americans.

By comparing classic machine learning classifiers to the PSI, a high-validated robust clinical decision tool, we have demonstrated the potential of machine learning as a predicting tool for severity indices. Application of machine learning to other conditions, where robust clinical decision tools are not yet available, may be of value.

One of the limitations of our study is the potential bias existing within the dataset. Comparing to the US census data [30], the dataset used for this study has more Caucasian population (81.28% vs 76.3% (census)). Black and Asian, on the other hand, are less populated (Black-8.5% vs 13.4% (census), Asian-1.44% vs 5.9% (census)). There could be biased based on the facilities where this dataset covers. The findings presented in this paper are limited to the characteristics of the dataset we inherited. The minority groups in the dataset have low number of cases and it’s difficult to detect differential patterns or correlating factors in those racial groups, if exist. Further studies are needed to understand the performance of machine learning and PSI across different racial groups and identify the correlating factors for each racial group.

## Data Availability

All data produced in the present study are available upon reasonable request to Regenstrief Center for Healthcare Engineering at Purdue University

## ACKNOWLEDGEMENT

We would like to thank the Regenstrief Center for Healthcare Engineering at Purdue University for providing the dataset.

